# Covid-19 and post-acute sick leave: a hybrid register and questionnaire study in the adult Danish population

**DOI:** 10.1101/2023.03.31.23288004

**Authors:** Elisabeth O’Regan, Ingrid Bech Svalgaard, Anna Irene Vedel Sørensen, Lampros Spiliopoulos, Peter Bager, Nete Munk Nielsen, Jørgen Vinsløv Hansen, Anders Koch, Steen Ethelberg, Anders Hviid

## Abstract

Long covid follows 10-20% of first-time SARS-CoV-2 infections, but the societal burden of long covid and risk factors for the condition are not well-understood. Here, we report findings about self-reported sick leave and risk factors thereof from a hybrid survey and register study, which included 37,482 RT-PCR confirmed SARS-CoV-2 cases and 51,336 test-negative controls who were tested during the index and alpha waves. An additional 33 individuals per 1000 took substantial sick leave following acute infection compared to persons with no known history of infection, where substantial sick leave was defined as >1 month of sick leave within the period 1-9 months after the RT-PCR test date. Being female, ≥50 years, and having certain pre-existing conditions such as fibromyalgia increased risks for taking substantial sick leave. Further research exploring this heterogeneity is urgently needed and may provide important evidence for more targeted preventative strategies.

## Introduction

Long covid has been termed to describe the persistence of wide-ranging, post-acute symptoms in individuals previously infected with SARS-CoV-2 (1). It has been estimated that 10-20% of people infected with SARS-CoV-2 develop long covid (2). With studies pointing to varying symptom clusters (3–6), unclear pathogenesis (1, 7), and a limited understanding of risk factors (8–11), long covid remains enigmatic. In particular, it is still unclear how many previously infected suffer from long covid to such an extent that it is severely disruptive to daily living and quality of life, and in turn also disruptive to society (12).

In Denmark, an earlier study on post-acute symptoms identified sick leave as a potential indicator of the burden of long covid symptoms, where both full- and part-time sick leave were more frequent among test-positives for SARS-CoV-2 compared to test-negatives (13). Still, the extent to which long covid symptoms translate to working ability remains understudied, and current literature on the subject has largely lacked control groups with no history of SARS-CoV-2 infection (5, 12, 14–17).

As many people have been infected and continue to be infected with SARS-CoV-2, further research is needed to understand the individual- and societal burden of long covid. Moreover, it is important to investigate the degree to which this burden is impacted by individual risk factors, such as age, sex, and preexisting health conditions. Deeper understanding of these risk factors can help guide clinical practice and target preventative public health measures.

The aim of this study was to evaluate the association between Covid-19 and post-acute sick leave and explore the possible impact of age, sex, and preexisting health conditions on this association.

## Methods

### Study context

#### Denmark’s universal SARS-CoV-2 testing strategy

In this study, it is important to consider Denmark’s national SARS-CoV-2 testing strategy and how this has facilitated the conduct of population-level studies with test-negative control groups, particularly before Omicron and the availability of home rapid antigen tests. In Denmark, universal testing for SARS-CoV-2 was implemented from the end of May 2020 and continued throughout the testing period under study from November 2020 to February 2021. Reverse transcription polymerase chain reaction (RT-PCR) tests were available and accessible for all adults free of charge and independent of the indication for acquiring a test (18). Additionally, persons admitted to hospitals were tested for SARS-CoV-2. During the testing period under study between November 2020 and February 2021 (index and alpha waves), the weekly PCR test incidence in Denmark ranged from 7,900 tests - 14,700 tests per 100,000 inhabitants (mean of 10,800 tests per 100,000 inhabitants) (19). Since February 2020, all RT-PCR test results have been registered in the Danish microbiology database (MiBa) (20).

#### Sick leave in Denmark

Most employees in Denmark are covered under the Act on Salaried Employees and are thus entitled to full pay during sick leave (21). Employers typically cover the first four weeks of sick leave, after which municipalities bear the costs.

### Study design

In this cohort study, we merged nationwide survey- and register data. In Denmark, all residents are assigned a unique identifier (the CPR-number) in the Danish Civil Registration System, and this number is commonly used to link individual-level data from varying data sources. A summary of the characteristics of our study population and which variables we used from survey responses and register data are presented in Figure 1.

**Figure 1:**
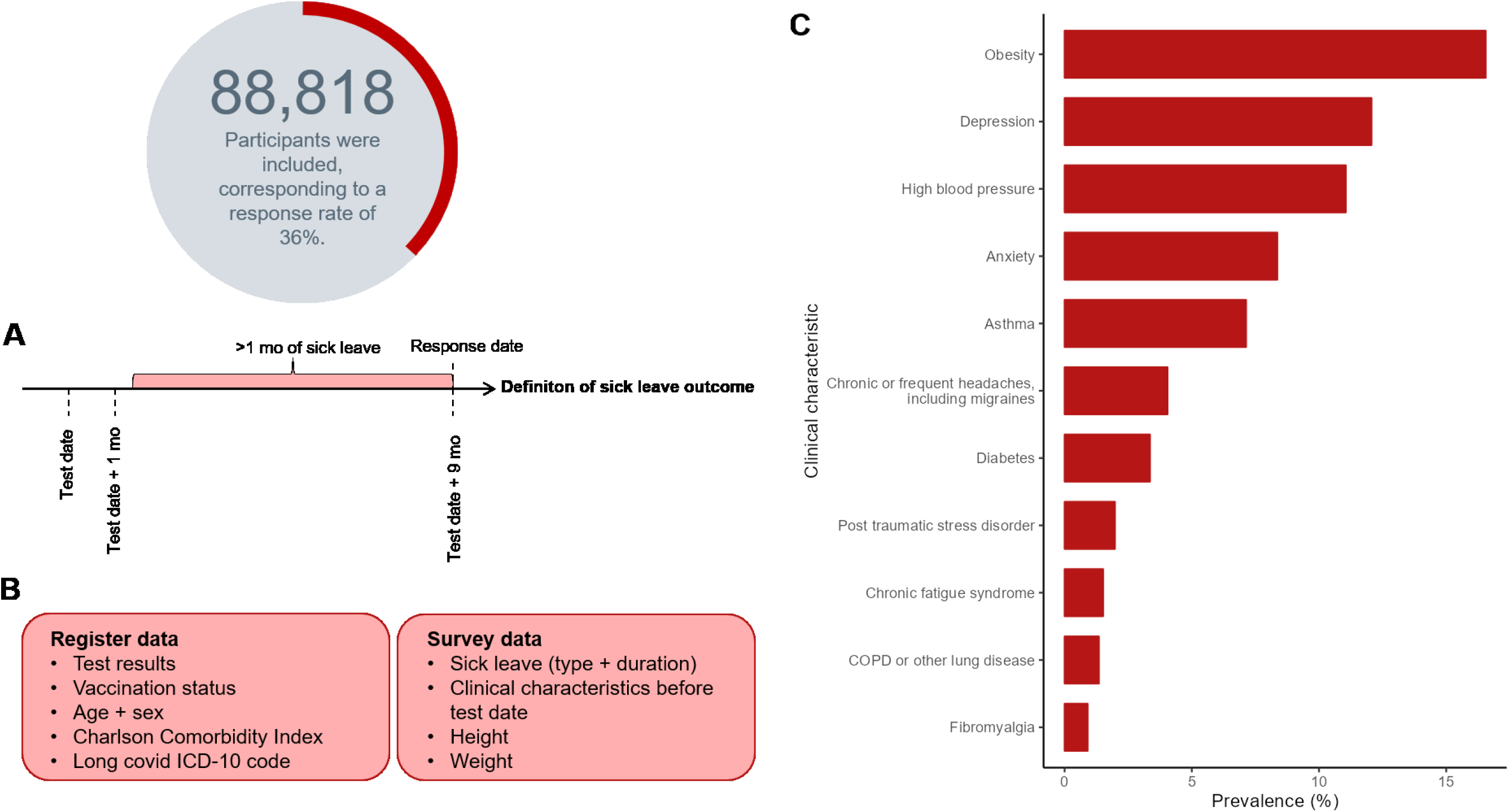
Overview of study population and study variables. N = 88,818 participants ages 15-65 were included (response rate = 36%). **Panel A:** Definition of the sick leave outcome, substantial sick leave. Substantial sick leave was defined as >1 month of sick leave in the period 1-9 months after the test. **Panel B:** Overview of study variables pulled from national register data and survey data. **Panel C:** Prevalence of each clinical characteristic/pre-existing condition in the total study sample, including both test-positives and -negatives for SARS-CoV-2.

#### The EFTER-COVID survey

To investigate self-reported sick leave after infection with SARS-CoV-2, we used data from a nationwide Danish survey, EFTER-COVID (AFTER COVID). This survey was launched in August 2021 to investigate the general health of the Danish population during the pandemic with a particular focus on long covid. Based on RT-PCR test results recorded in MiBa, individuals were sent an invitation to participate in the study via the national digital mail system. This system (“e-Boks”) enables secure electronic communication with public authorities and is used by over 90% of all Danish residents aged ≥15 years (22). EFTER-COVID survey data were collected using Danish- or English-language web-based questionnaires created in SurveyXact (23), which could be filled out using a PC, tablet, or smartphone.

All Danish residents who had an e-Boks account and obtained a first positive RT-PCR test result for SARS-CoV-2 registered in MiBa during the period from November 2020 to February 2021 were invited to participate in the EFTER-COVID survey. Additionally, test-negative controls were randomly selected using incidence density sampling on the test date with a ratio of 2:3 between test-positive and -negative persons. This ratio was chosen to compensate for a lower expected response rate among controls compared to cases. Importantly, the test-negative controls did not have a registered positive test result in MiBa at any time point prior to receiving or responding to the questionnaire, which we could reassure due to the extensive national testing strategy. In this study, we included participants who responded to a retrospective questionnaire 9 months after their test date. Other studies which have used the EFTER-COVID survey data (13, 24, 25) include participants who responded at other points in time after testing and thus reflect different subsets of EFTER-COVID data, which contain more than 840,000 survey participants (26).

#### Data sources

Using the CPR-number, we enriched the EFTER-COVID questionnaire data with register-based information on age and sex, Covid-19 vaccinations from the Danish Vaccination Register (DDV), SARS-CoV-2 test results and (re)infection history registered in MiBa, and comorbidities from the Danish National Patient Register (DNPR) five years prior to each participant’s test date.

The questionnaire included questions about baseline characteristics of the participant, including height, weight, education, smoking habits, alcohol consumption, and health conditions preceding the individual’s test date. In addition, participants were asked about the amount of full- or part-time sick leave they took between their test date (indicated in the questionnaire), and the day they responded to the survey. Test-negatives were asked whether they suspected ever having had Covid-19, e.g., if they had received a seropositive test result. All questions required a response to complete the questionnaire, except for height, weight, smoking and alcohol consumption.

From the DNPR, we obtained information on in- and outpatient diagnoses coded using the 10^th^ revision of the International Statistical Classification of Diseases and Related Problems (ICD-10), which enabled the calculation of Charlson Comorbidity Index. We also extracted information on the long covid ICD-10 code (ICD-10 code B948A) from the DNPR. A complete description of how we categorized these variables can be found in Table S1.

#### Exclusion criteria

Participants who did not complete the questionnaire were excluded. Furthermore, we did not include individuals who indicated that they believed they previously had SARS-CoV-2 due to receiving a seropositive result for SARS-CoV-2. Participants who were >65 years were also excluded due to retirement age, where age was calculated on the test date. See Figure S1 for a detailed flowchart of our inclusion and exclusion criteria.

#### Outcomes

All participants, regardless of test status, were asked whether they took sick leave around the time of their test date or at any time point after the test. Individuals who responded “yes” to taking sick leave more than four weeks after the test were then asked whether their sick leave was full-time, part-time, or both and for how long they were on full- and/or part-time sick leave.

A binary outcome variable was defined as having taken “*no or ≤4 weeks of full-time sick leave >4 weeks after the test date*” or “*>4 weeks of full-time sick leave >4 weeks after the test date*”. The latter was treated as indicative of substantial sick leave, i.e., at least one full month of sick leave was taken over an eight-month period one month after the test. An identical outcome was defined for part-time sick leave.

We chose to examine sick leave from one month after the test to capture sick leave after the acute infection period among test-positives. The self-reported sick leave was not necessarily taken consecutively such that it could capture multiple periods of sick leave due to fluctuating symptoms.

Participants who reported taking only sick leave during the week up to the test and up to four weeks after the test where included in the category “*no or <4 weeks of sick leave >4 weeks after the test date*”. For an illustration of how we defined our sick leave outcome, see Figure 1, Panel A.

### Statistical methods

The prevalence of substantial sick leave among test-positives and -negatives were compared using risk differences (RDs), which give the difference between the risk of an outcome in the exposed group and the unexposed group. Parametric g-computation on logistic regression (27) was used to estimates RDs with 95% confidence intervals obtained using bootstrap random resampling with 1000 iterations comparing test-positive and -negative individuals with adjustments for age, sex, Charlson Comorbidity Index, and education level. RDs for the risk of substantial sick leave are expressed in percentage points. *P*-values for the association between test status and the survey participant characteristics were estimated using student’s t-test for continuous variables and Pearson’s Chi-squared test for categorical variables.

To investigate possible risk groups for substantial sick leave following infection with SARS-CoV-2, we conducted analyses on sub-populations defined by possible risk factors. These risk factors were defined apriori based on available variables in the survey. These included middle to older age (categorized as ≥50 years), female sex, obesity, diabetes, asthma, high blood pressure, COPD or other chronic lung disease, chronic or frequent headaches/migraines, and the following health conditions diagnosed by a medical doctor before the test: depression, anxiety, post-traumatic stress disorder, chronic fatigue syndrome, and fibromyalgia. In addition, we restricted analyses by educational level and healthcare workers. RDs with 95% confidence intervals were estimated as described above.

All statistical analyses were carried out in R version 4.1.3 (28). The R-packages “riskCommunicator” (29) was used for modelling and “forestploter” (30) for data visualization (forest plots).

## Results

### Overview of study population

Out of 294,035 invited, a total of 106,917 persons fully completed a questionnaire nine months after testing positive or negative for SARS-CoV-2 (response rate 36.4%). After all exclusion criteria were applied, the study population consisted of 88,818 individuals, of which 37,482 had had SARS-CoV-2 infection confirmed with a positive reverse transcription polymerase chain reaction (RT-PCR) test (Figure S1). Of all participants, 64.3% were female, and the mean age was 45 years (SD 13.8) (Table 1). Less than 1% of participants had received one or more doses of a vaccine against Covid-19. Based on self-reported height and weight, 16.6% of the population was identified as obese. Apart from obesity, the most frequent preexisting clinical characteristics were depression, high blood pressure, and anxiety (Table 1, Figure 1, Panel C).

**Table 1:**
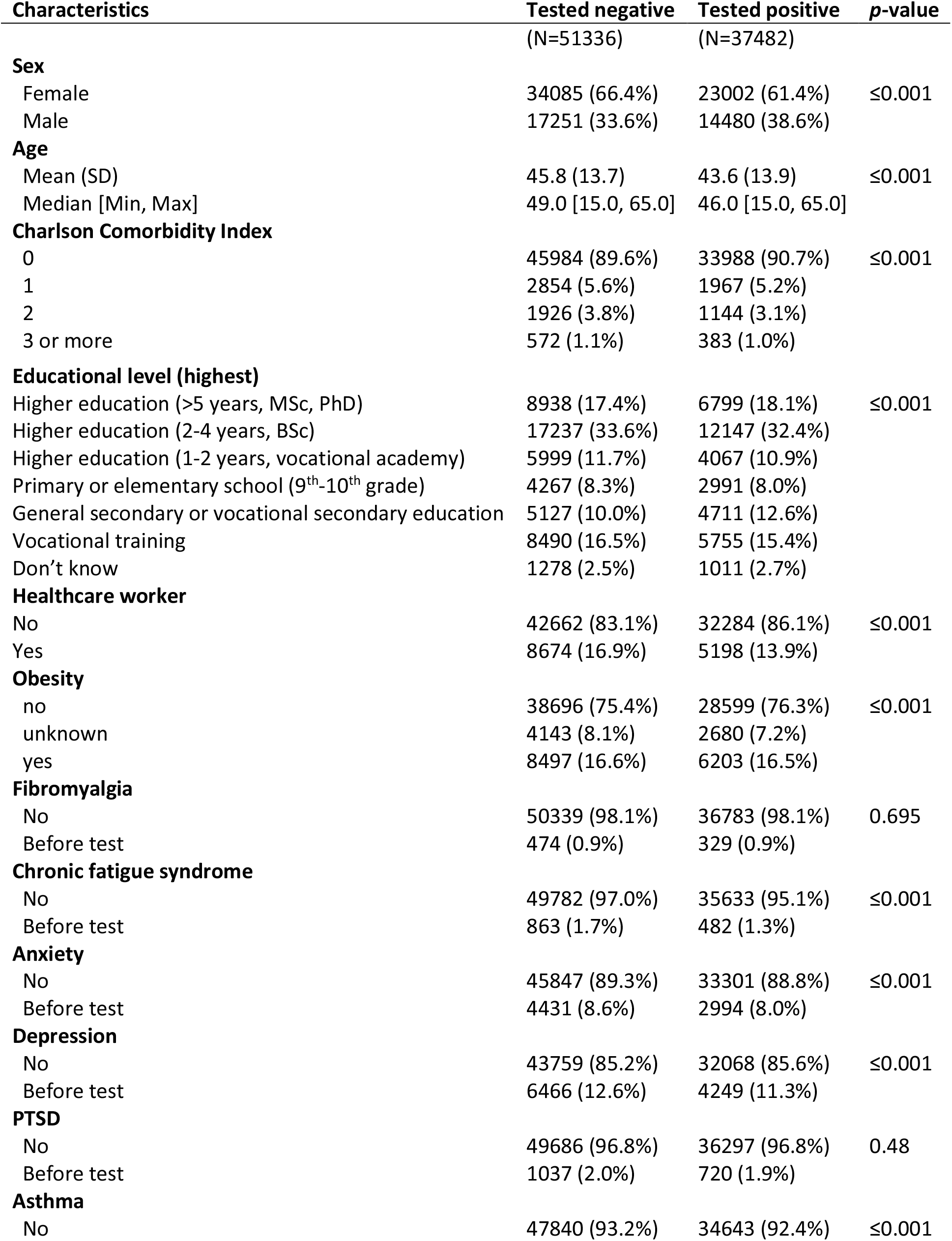
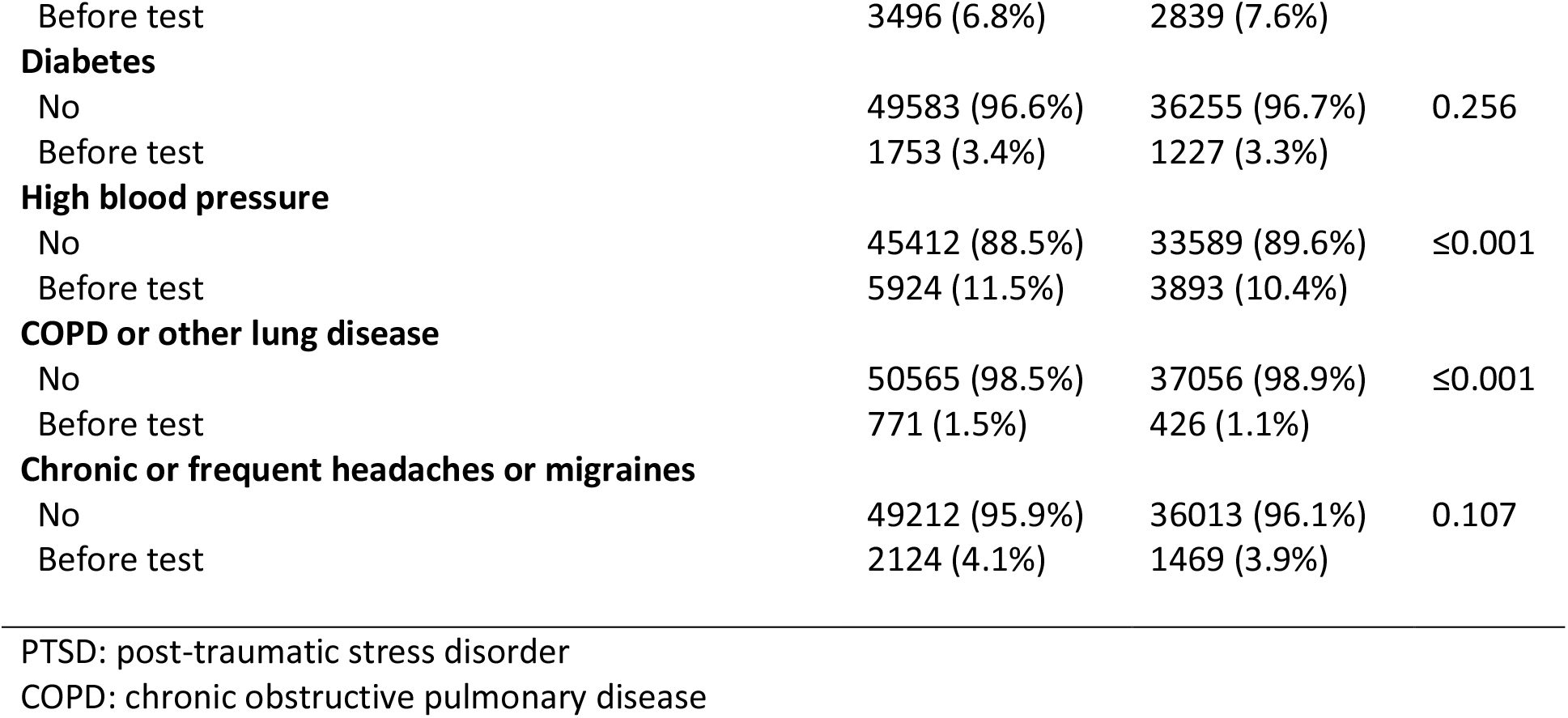
Characteristics of 88,818 study participants who obtained a positive or negative PCR test for SARS- CoV-2.

| Characteristics | Tested negative<br>(N=51336) | Tested positive<br>(N=37482) | p-value |
| --- | --- | --- | --- |
| **Sex** |  |  |  |
| Female | 34085 (66.4%) | 23002 (61.4%) | ≤0.001 |
| Male | 17251 (33.6%) | 14480 (38.6%) |  |
| **Age** |  |  |  |
| Mean (SD) | 45.8 (13.7) | 43.6 (13.9) | ≤0.001 |
| Median [Min, Max] | 49.0 [15.0, 65.0] | 46.0 [15.0, 65.0] |  |
| **Charlson Comorbidity Index** |  |  |  |
| 0 | 45984 (89.6%) | 33988 (90.7%) | ≤0.001 |
| 1 | 2854 (5.6%) | 1967 (5.2%) |  |
| 2 | 1926 (3.8%) | 1144 (3.1%) |  |
| 3 or more | 572 (1.1%) | 383 (1.0%) |  |
| **Educational level (highest)** |  |  |  |
| Higher education (>5 years, MSc, PhD) | 8938 (17.4%) | 6799 (18.1%) | ≤0.001 |
| Higher education (2-4 years, BSc) | 17237 (33.6%) | 12147 (32.4%) |  |
| Higher education (1-2 years, vocational academy) | 5999 (11.7%) | 4067 (10.9%) |  |
| Primary or elementary school (9<sup>th</sup>-10<sup>th</sup> grade) | 4267 (8.3%) | 2991 (8.0%) |  |
| General secondary or vocational secondary education | 5127 (10.0%) | 4711 (12.6%) |  |
| Vocational training | 8490 (16.5%) | 5755 (15.4%) |  |
| Don't know | 1278 (2.5%) | 1011 (2.7%) |  |
| **Healthcare worker** |  |  |  |
| No | 42662 (83.1%) | 32284 (86.1%) | ≤0.001 |
| Yes | 8674 (16.9%) | 5198 (13.9%) |  |
| **Obesity** |  |  |  |
| no | 38696 (75.4%) | 28599 (76.3%) | ≤0.001 |
| unknown | 4143 (8.1%) | 2680 (7.2%) |  |
| yes | 8497 (16.6%) | 6203 (16.5%) |  |
| **Fibromyalgia** |  |  |  |
| No | 50339 (98.1%) | 36783 (98.1%) | 0.695 |
| Before test | 474 (0.9%) | 329 (0.9%) |  |
| **Chronic fatigue syndrome** |  |  |  |
| No | 49782 (97.0%) | 35633 (95.1%) | ≤0.001 |
| Before test | 863 (1.7%) | 482 (1.3%) |  |
| **Anxiety** |  |  |  |
| No | 45847 (89.3%) | 33301 (88.8%) | ≤0.001 |
| Before test | 4431 (8.6%) | 2994 (8.0%) |  |
| **Depression** |  |  |  |
| No | 43759 (85.2%) | 32068 (85.6%) | ≤0.001 |
| Before test | 6466 (12.6%) | 4249 (11.3%) |  |
| **PTSD** |  |  |  |
| No | 49686 (96.8%) | 36297 (96.8%) | 0.48 |
| Before test | 1037 (2.0%) | 720 (1.9%) |  |
| **Asthma** |  |  |  |
| No | 47840 (93.2%) | 34643 (92.4%) | ≤0.001 |
| Before test | 3496 (6.8%) | 2839 (7.6%) |  |
| **Diabetes** |  |  |  |
| No | 49583 (96.6%) | 36255 (96.7%) | 0.256 |
| Before test | 1753 (3.4%) | 1227 (3.3%) |  |
| **High blood pressure** |  |  |  |
| No | 45412 (88.5%) | 33589 (89.6%) | ≤0.001 |
| Before test | 5924 (11.5%) | 3893 (10.4%) |  |
| **COPD or other lung disease** |  |  |  |
| No | 50565 (98.5%) | 37056 (98.9%) | ≤0.001 |
| Before test | 771 (1.5%) | 426 (1.1%) |  |
| **Chronic or frequent headaches or migraines** |  |  |  |
| No | 49212 (95.9%) | 36013 (96.1%) | 0.107 |
| Before test | 2124 (4.1%) | 1469 (3.9%) |  |

PTSD: post-traumatic stress disorder
COPD: chronic obstructive pulmonary disease

### Prevalence of substantial sick leave

The prevalence of substantial sick leave was 1.4% among test-negatives compared to 4.5% among test-positives. Among test-negatives only, the prevalence of substantial sick leave was similar across age groups. Conversely, for test-positives, the prevalence of substantial sick leave increased with age (Figure 2, Panel C). Notably, out of all test-positives who took substantial sick leave, 354 individuals (21.1%) had received a diagnosis with sequelae of SARS-CoV-2 (ICD-10 code B948A) (Figure 2, Panel A) compared to only 556 (1.6%) of the test-positives who did *not* have substantial sick leave.

**Figure 2:**
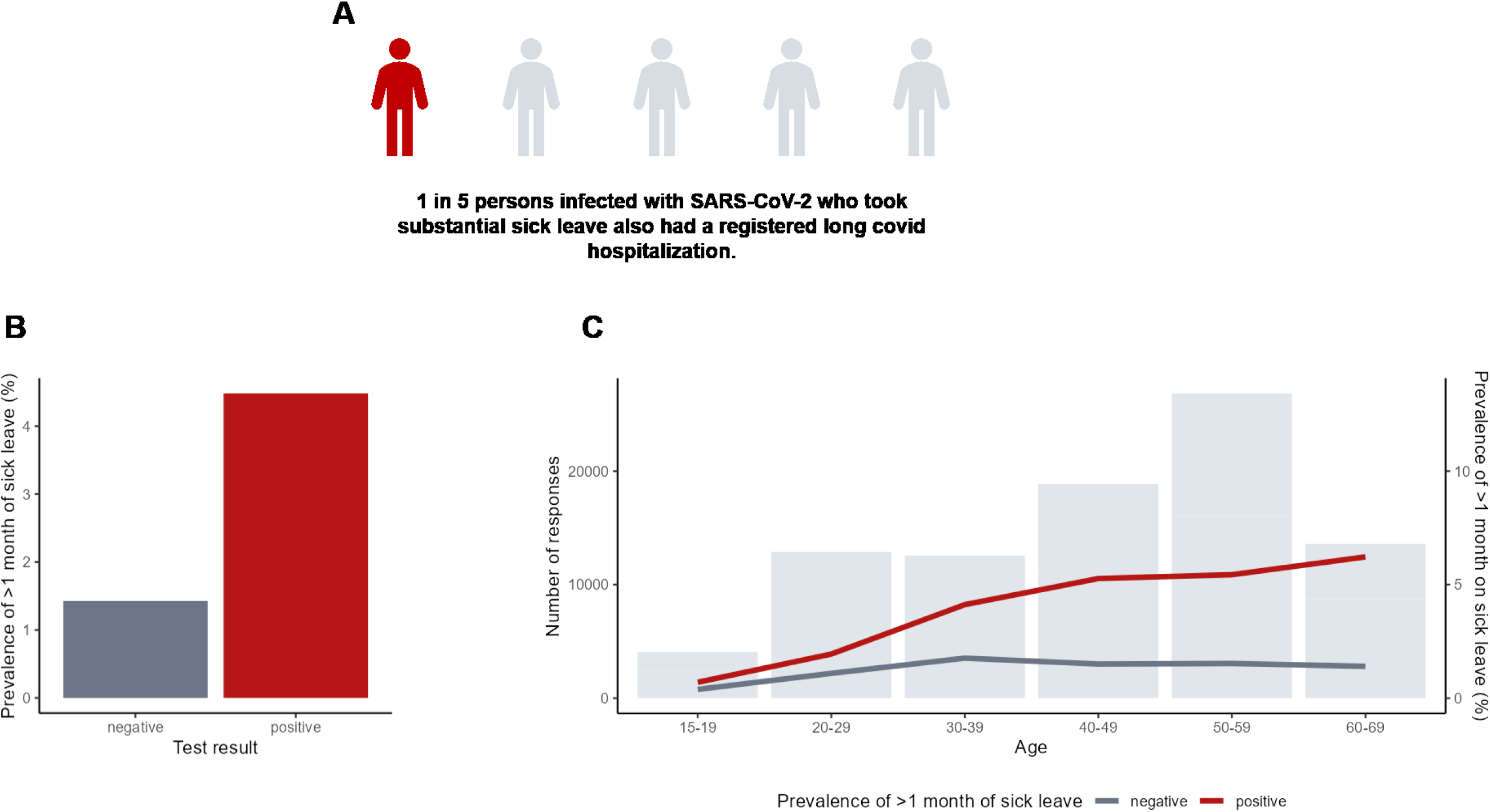
Prevalence of substantial sick leave, defined as >1 month of sick leave in the period 1-9 months after the test date. N = 88,818 participants ages 15-65 were included (response rate = 36%). N_positive_ = 51,336, N_negative_ = 37,482. **Panel A:** 21.1% of persons infected with SARS-CoV-2 who took substantial sick leave also had a registered long covid hospitalization. Long covid hospitalization was defined as having a registered International Classification of Diseases 10^th^ Revision (ICD-10 code B948A) in the period 1-9 months after the test date and having no history of this diagnosis within the year prior to the test date. **Panel B:** Unadjusted prevalence of substantial sick leave by SARS-CoV-2 PCR test result. **Panel C:** Number of responses and prevalence of substantial sick leave by age group and PCR test result.

In view of possible risk groups identified in previous literature (1, 10, 14, 31) we stratified on sex, middle to older age (>50 years), and pre-existing health conditions. Across strata, the prevalence of substantial sick leave was higher among test-positives compared to test-negatives. Among persons with pre-existing conditions, the baseline prevalence of substantial sick leave, i.e., among test-negatives, was higher than for the test-negative general population (1.4%), and the highest background prevalence was among individuals with chronic fatigue syndrome (4.1%) (Figure 3).

**Figure 3:**
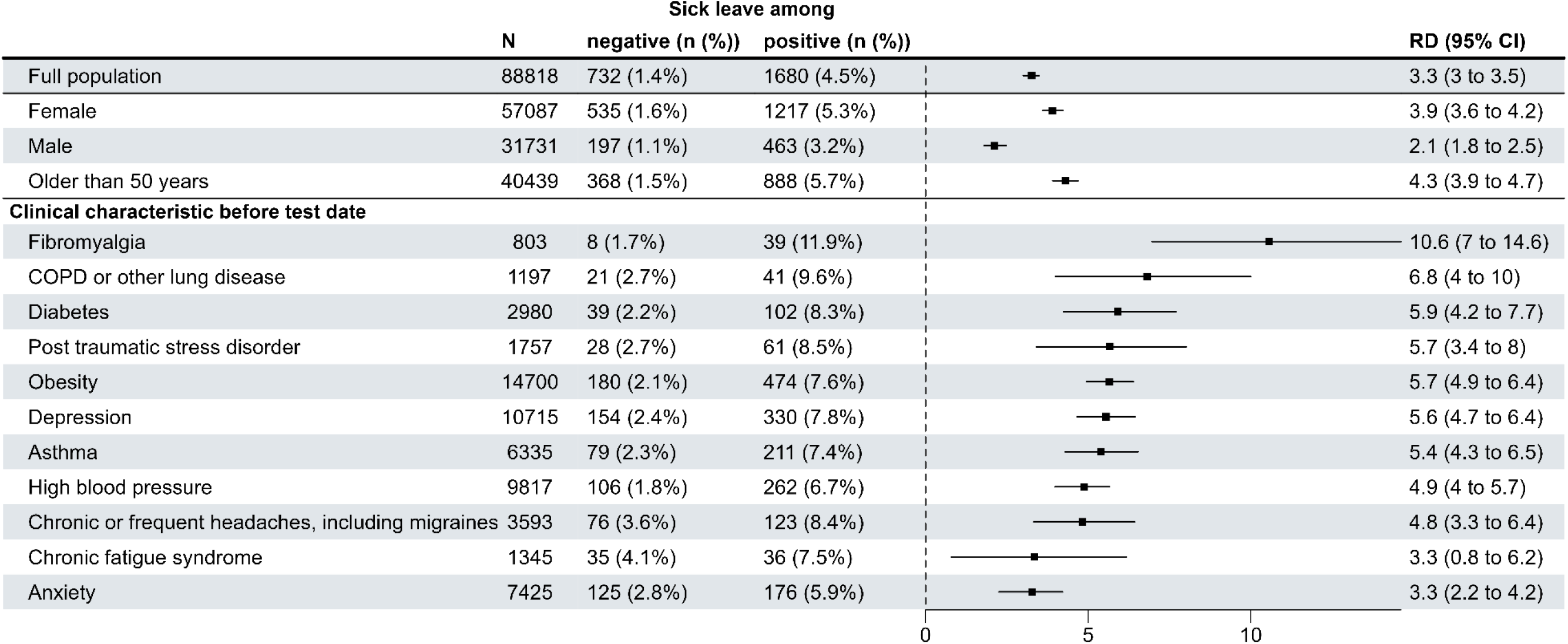
Risk differences (RDs) and 95% confidence intervals (CI) for full-time substantial sick leave taken one to nine months after the test date between SARS- CoV-2 test-positives and test-negatives for the total study population and possible long covid risk groups. RDs are adjusted for age, sex, Charlson Comorbidity Index, educational level, and select pre-existing conditions (chronic diseases). N = 88,818 participants ages 15-65 years were included (response rate = 36%). N_positive_ = 51,336, N_negative_ = 37,482. RDs are expressed in percentage points.

### Risk differences for substantial sick leave

Nine months after testing for SARS-CoV-2 during the index and alpha waves, persons infected with SARS-CoV-2 had a higher risk (RD 3.3, 95% CI 3.0 to 3.5) of taking substantial sick leave after their acute infection (>1 month of sick leave within 1-9 months after the test date) compared to test-negatives with no known history of SARS-CoV-2 infection (Figure 3). Changing the definition of substantial sick leave by increasing the duration resulted in attenuation of the RD (e.g., RD = 0.5, 95% CI 0.4 to 0.6) for substantial sick leave defined as at least 6 months (Figure S3). A distributional plot underlying substantial sick leave by age (<50 years, ≥50 years) and sex is available in Figure S4.

Risk differences were greater for females (RD 3.9, 95% CI 3.6 to 4.2) than males (RD 2.1, 95% CI 1.8 to 2.5). For middle-older age (≥50 years) and all pre-existing conditions except chronic fatigue syndrome and anxiety, estimated RDs were higher than that in the general population. The largest risk differences were observed for persons with fibromyalgia (RD 10.6, 95% CI 7 to 14.6), COPD or other lung disease (RD 6.8, 95% CI 4 to 10) and diabetes (RD 5.9, 95% CI 4.2 to 7.7). Obesity, the most frequent clinical characteristic, had a larger risk difference (RD 5.7, 95% CI 4.9 to 6.4) than that in the general population (RD 3.3, 95% CI 3 to 3.5) (Figure 3). RDs by education level showed that individuals with a “higher education of 2-4 years (e.g., nurse, preschool teacher, bachelor of engineering)” had a larger risk difference (RD 3.8, 95% CI 3.4 to 4.2) than that in individuals with a “higher education of ≥5 years (e.g., master’s degree or PhD)” (RD 1.9, 95% CI 1.4 to 2.4) (Table S2). Healthcare workers (N=13,872) also had a larger risk difference (RD 4.6, 95% CI 3.9 to 5.3) than the general population.

### Risk differences for part-time sick leave

Furthermore, persons infected with SARS-CoV-2 had a higher risk (RD 2.1, 95% CI 2.0 to 2.3) of substantial part-time sick leave compared to test-negatives. Similar results when stratifying on sex were observed for both part-time and full-time sick leave. However, with regards to individuals ≥50 years, a higher risk was noted for full-time sick leave than for part-time sick leave. For full-time sick leave, this age group had a higher RD (RD 4.3, 95% CI 3.9 to 4.7) than that of the full population (RD 3.3, 95% CI 3.0 to 3.5). For part-time sick leave, individuals ≥50 years had a similar risk (RD 2.3, 95% CI 2.1 to 2.6) compared to that of the full study population (RD 2.1, 95% CI 2.0 to 2.3) (Figure S2).

## Discussion

### Key findings

In this study, we used sick leave as a proxy for long covid and explored sick leave among a range of possible long covid risk groups. We found that individuals infected with SARS-CoV-2 during the index and alpha waves had a significantly higher risk of substantial sick leave (3.3 percentage point increase in risk), where substantial sick leave was defined as >1 month of self-reported sick leave within the period 1-9 months after the test. Notably, 20.1% of the test-positives who took substantial sick leave also had a registered ICD-10 code for long covid. Patients who receive a long covid diagnosis represent only a small proportion of individuals suffering from post-acute symptoms; as such, the fact that one in five of those experiencing substantial sick leave also have a long covid diagnosis supports the validity of using substantial sick leave as a proxy for long covid burden. We identified females, persons ≥ 50 years, obesity and a variety of pre-existing health conditions as possible risk factors, where the risk difference between testing positive and negative was greater than that of the overall study population.

### Other risk factors

Other work has examined sick leave in persons previously infected with SARS-CoV-2; however, differences in risk measures, timing of measurements, national testing strategies, and definitions of sick leave vary between studies and should be carefully considered. A Danish register-based cohort study with 7,466 participants examined return-to-work following first-time infections occurring between January and May 2020, where 81.9% returned to work within four weeks of their first positive SARS-CoV-2 test (32). Although this study did not have a test-negative control group, the authors examined the cumulative incidence of return to work between patients admitted to hospitals with SARS-CoV-2 and patients admitted with influenza, which suggested that patients with SARS-CoV-2 had a reduced chance of returning to work compared to patients admitted with influenza. Additionally, a Danish cross-sectional study examined the influence of long covid on activities of daily living among 448 long covid patients reported that 56% needed sick leave and 94% were referred to rehabilitation (17). In a German register-based study with 30,950 individuals diagnosed with SARS-CoV-2, a reported 5.8% of individuals took more than four weeks of sick leave between March 2020 and February 2021 (14). These studies, together with ours, point to sick leave as a suitable indicator of the personal and societal burden of long covid. Intriguingly, what may be more telling is the heterogeneity of sick leave following SARS-CoV-2 for the purpose of identifying long covid risk groups.

Female sex has been identified as a possible risk factor for long covid (7). Some studies have indicated that females needed longer sick leave following infection with SARS-CoV-2 than their male counterparts (14, 32). Others have pointed to interactions between age and sex, e.g., less sick leave in infected females ages 20-44 compared to infected females ages 45-70 years (33). Additionally, a Swedish registry-based cohort study reported that people with recurrent sick leave were older, more often female, and more likely to have been on sick leave prior to the pandemic (16). In our study, we observed that the risk difference for substantial sick leave between SARS-CoV-2 test-positives and -negatives was greater for females (RD 3.9, 95% CI 3.6 to 4.2) than for males (RD 2.1, 95% CI 1.8 to 2.5), and that females as well as persons ≥50 years had a slightly higher prevalence of sick leave compared to the general population, irrespective of test status. Importantly, severity of SARS-CoV-2 infection is associated with increased age, pre-existing medical conditions, and male sex (34), and severity of infection has been associated with long covid (7). It is therefore unclear as to why females appear to be a risk group for substantial sick leave following acute SARS-CoV-2 infection but males are not. Further research examining age-sex interactions and the impact of multimorbidity on long covid outcomes are needed (35).

A German register-based study by Jacob et al. (14) also explored a range of possible risk factors, some of which overlapped with risk factors that we explored. As in our study, the authors also found that diabetes and high blood pressure were positively and significantly associated with long-term sick leave. However, in contrast to our study findings, the authors did not find obesity and asthma to increase the risk of long-term sick leave. Furthermore, the authors reported that anxiety and somatoform disorders were not associated with the risk of longer-term sick leave. Similarly, in our study, neither anxiety nor chronic fatigue syndrome, which is categorized as a somatoform disorder by some, increased the risk of substantial sick leave; however, new-onset chronic fatigue syndrome is a known complication of long covid (7).

Fibromyalgia, COPD/other chronic lung diseases, and obesity have also been cited as possible risk factors for long covid in other work. A US study (preprint) (N=89,843) found that long covid patients were more likely to have a history of fibromyalgia (OR 2.3, 95% CI 1.3 to 3.8) and chronic pulmonary lung disease (OR 1.9, 95% CI 1.5 to 2.6) compared to matched test-positive controls without long covid (36). Additionally, obesity has been identified as a risk factor for severe acute SARS-CoV-2 infection (1), and inpatient care following infection has been suggested to predict longer sick leave (31).

### Industry

Finally, individuals’ line of work can impact the need for long covid-related sick leave. Although we were not able to examine most individuals’ professions, we did observe that individuals with a higher education of 2-4 years had a larger risk difference of taking substantial sick leave compared to individuals with a postgraduate education. We posit that these differences could be attributed to job adaptability to work-from-home and self-pacing, i.e., persons with a post-graduate education may work desk jobs whereas persons with a higher education of 2-4 years (e.g., nurse, preschool teacher, bachelor of engineering) may need to be on-site. Similar findings were reported in a Norwegian study, which estimated the industry-specific impact of the Omicron wave on sick leave compared to corresponding months from 2017-2020 (37). Persons within the food and accommodation industry had the highest increase in sick leave (4.4 percentage points increase, 95% CI 4.3-4.5), suggesting that individuals with jobs not-suited for work from home required more sick leave.

### Strengths and limitations

The present study describes population-level data on post-acute sick leave as a possible consequence of infection with SARS-CoV-2. In contrast to most existing literature on the subject of long covid and working ability, this study used a test-negative control group, allowing us to consider background prevalence of sick leave for the study population and among possible risk groups, including people with various conditions which preceded their SARS-CoV-2 test. Furthermore, our sick leave outcome variable captures fluctuating illness, which is useful given the irregularity of long covid symptoms.

We consider the main limitations of this study to be its self-reporting nature and potential participation and recall bias. One shortcoming of the self-reported sick leave variable is that the duration of sick leave cannot be directly attributed to long covid symptoms, as the questionnaire asked about sick leave in general rather than sick leave associated with SARS-CoV-2 infection. A possible alternative to studying self-reported sick leave was using register-based sick leave benefits. However, the prevalence of register-based sick leave benefits was previously examined for infections occurring between January and May 2020 in Denmark (32), where the authors used data from the Danish Register for Evaluation of Marginalization (DREAM). DREAM primarily covers sick leave benefits which are granted after 30 days of sickness absence. A disadvantage of using this data is that it largely excludes shorter periods of absence (38) and thus is not well-suited for capturing fluctuating illness as a possible consequence of long covid.

Participation bias may have occurred, where individuals living with poor health or long covid symptoms may have taken more interest in participating. Alternatively, some individuals living with long covid symptoms may have felt too poorly to participate. In addition, the retrospective study design is vulnerable to recall bias, where some participants may not remember how much sick leave they took over the 9 months following their test. We sought to reduce this by multiple choice between pre-defined sick leave durations instead of using free-text. While the RD was attenuated for increasing durations of sick leave as expected, the RD for at least six months (RD 0.5, 95% CI 0.4 to 0.6) is still quite striking considering the large number of infected globally. Lastly, our results capture index- and alpha-variant infections which largely occurred before SARS-CoV-2 vaccine rollout and we cannot exclude that the absolute magnitude of our results are attenuated by vaccinations and the omicron variant.

### Perspectives

Long covid is a concerning, contemporary public health issue with many unanswered questions, particularly regarding manifestations of symptom burden and risk factors. This study provides much-needed information on post-acute sick leave following SARS-CoV-2 infection in a general population and the risk of post-acute sick leave for people with comorbid conditions. The results from this study may be particularly useful to public health stakeholders in guiding evidence-based decisions concerning targeted preventative strategies. Further long covid research and funding initiatives are critical, particularly to increase knowledge of immunopathogenic mechanisms, phenotypes, risk factors, and the impact of different (sub)variants and SARS-CoV-2 vaccines.

## Conclusion

Among individuals infected with SARS-CoV-2 during the index and alpha waves, an additional 33 individuals per 1000 took substantial sick leave within 1-9 months following acute infection compared to persons with no known infection. Females, older individuals, and individuals with pre-existing conditions such as fibromyalgia, COPD/chronic lung diseases, and obesity were affected. This study may be used to help inform the healthcare and research workforce of the impact of long covid on working ability and to motivate improved diagnostic and treatment options.

## Data Availability

The datasets used in this study comprise sensitive, individual-level information from completed questionnaires and national register data. According to the Danish data protection legislation, the authors are not permitted to share these sensitive data directly upon request. However, the data are available for research purposes upon request to the Danish Health Authority (register data) and Statens Serum Institut (questionnaire data),  as well as within the framework of the Danish data protection legislation and any required permission from authorities. Data request processing can take an expected three to six months.

## Contributors

This study was designed and initiated by PB, SE, AH, AK, AS, IB, and EO. The questionnaire was designed by AS, PB, NN, AK, SE.

Collection of data including programming related to this manuscript AS, JH, PB, IM, and LS. Data analysis was carried out by IB.

The first draft was written by EO, IB. Supervision of the study was carried out by AH. All authors have critically revised the manuscript.

All authors have approved the final version of the manuscript and agreed to be accountable for all aspects of the work.

EO and IB contributed equally to this manuscript. The corresponding author attests that all listed authors meet authorship criteria and that no others meeting the criteria have been omitted. EO and AH are the guarantors.

## Ethics approval

This study was performed as a surveillance study as part of the governmental institution Statens Serum Institut’s (SSI) advisory tasks for the Danish Ministry of Health. SSI’s purpose is to monitor and fight the spread of disease in accordance with section 222 of the Danish Health Act. According to Danish law, national surveillance activities carried out by SSI do not require approval from an ethics committee.

Participation in the study was voluntary. The invitation letter to participants contained information about their rights under the Danish General Data Protection Regulation (rights to access data, rectification, deletion, restriction of processing and objection). After reading this information, it was considered informed consent if participants agreed and clicked on the link to fill in the questionnaires.

## Acknowledgements

The authors would like to thank all EFTER-COVID participants for making this research possible by responding to the questionnaires.

We would also like to thank the members of the EFTER-COVID stakeholder group, including representatives from universities, long-term sequelae clinics, and other relevant hospital departments or health institutions, for offering their feedback on the questionnaires and contributing to stakeholder meetings.

## Supplementary Material

**Figure S1:**
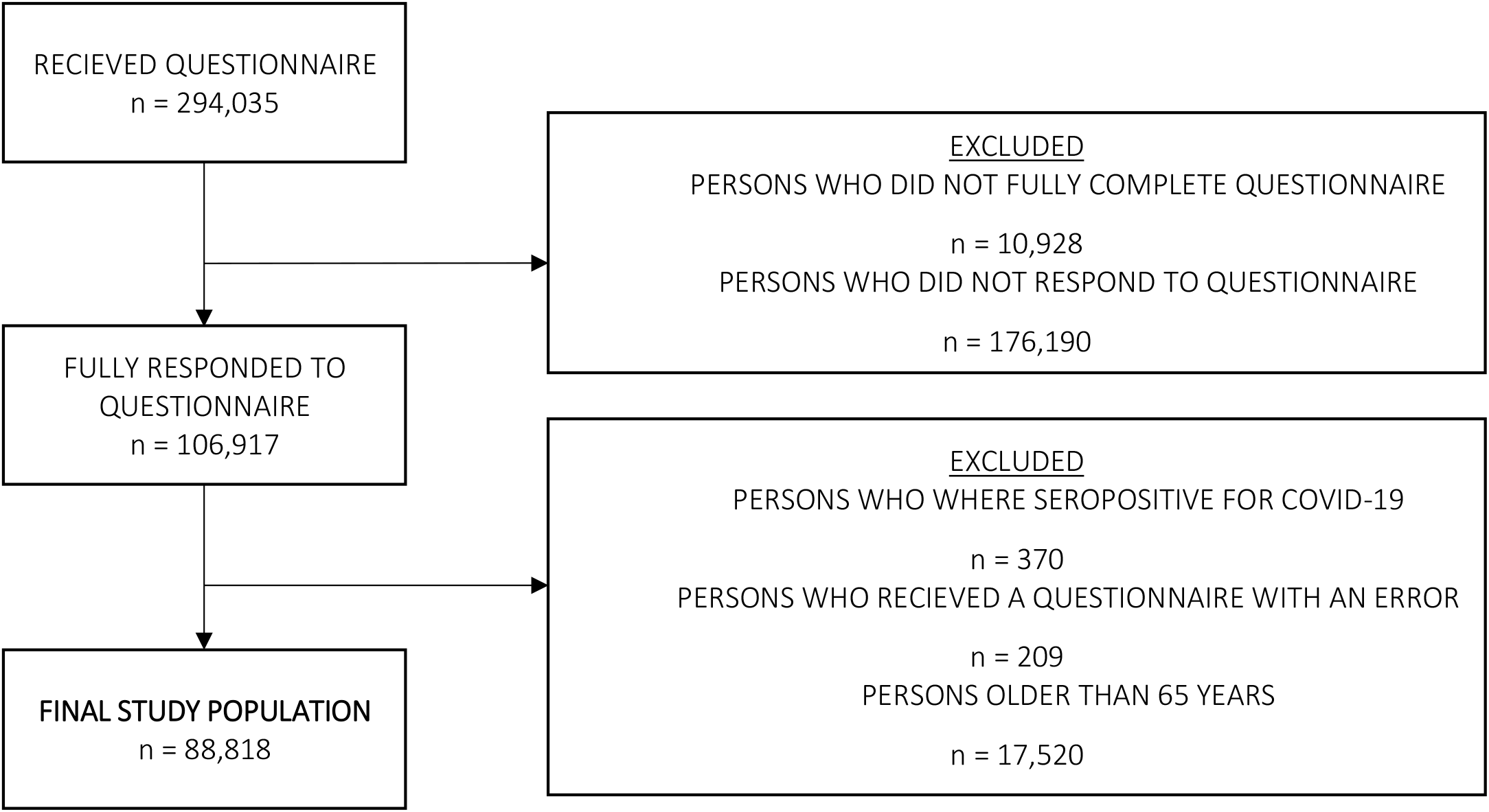
Flowchart of study population.

**Figure S2:**
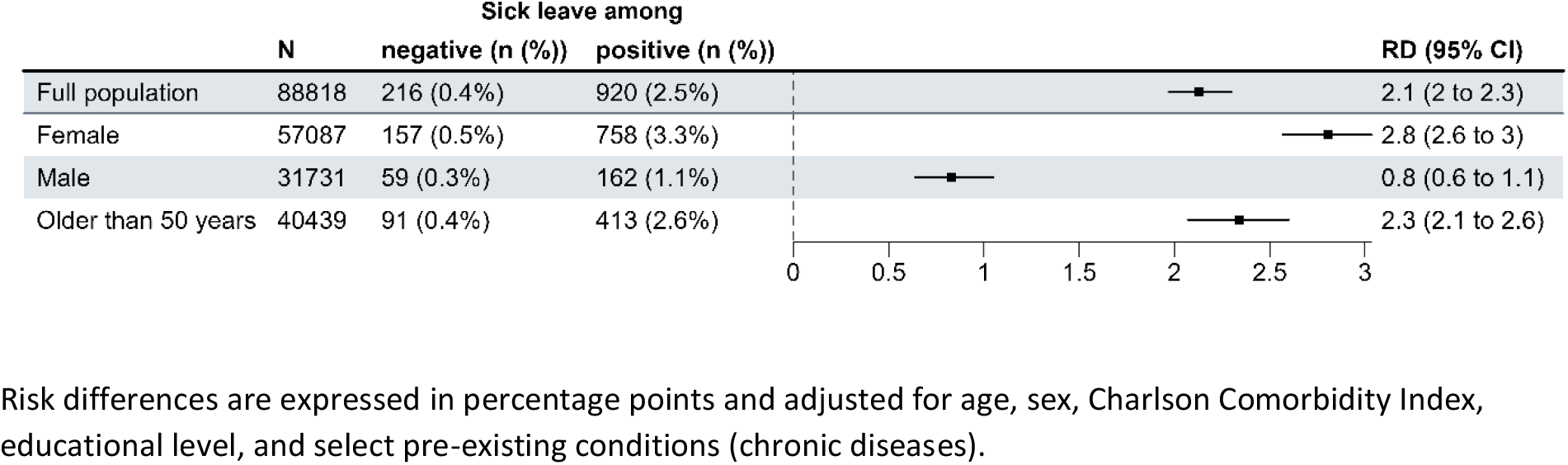
Risk differences (RDs) and 95% confidence intervals (CIs) for part-time substantial sick leave one to nine months after test date between SARS-CoV-2 test-positives and negatives.

**Figure S3:**
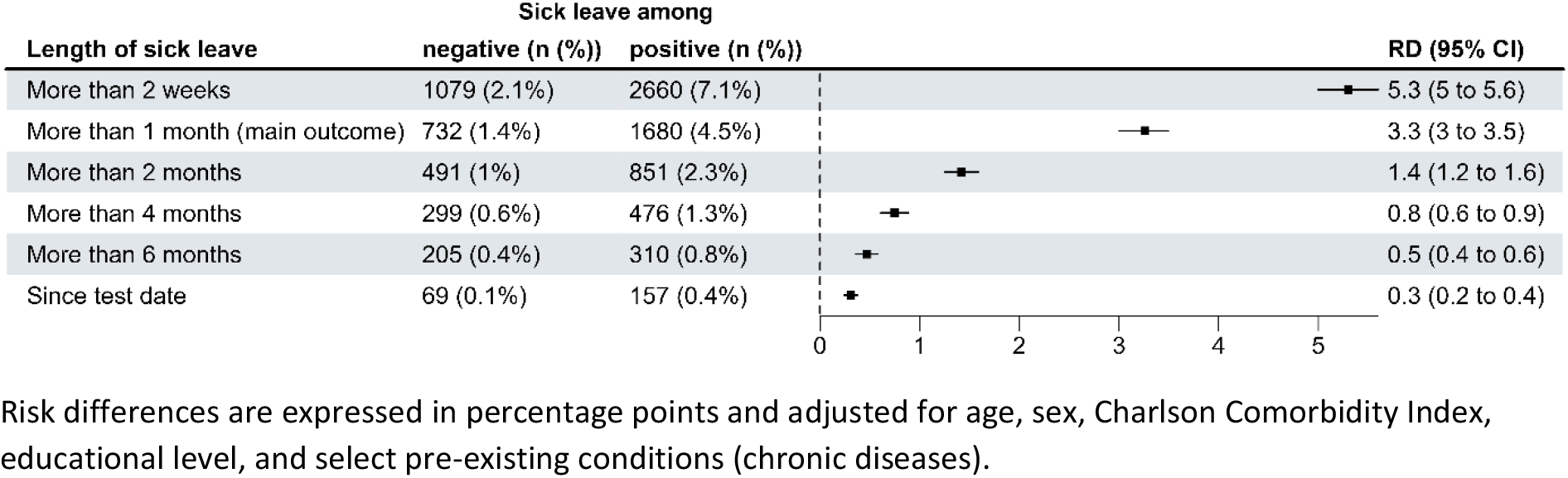
Risk differences (RDs) and 95% confidence intervals (CIs) for sick leave by differing duration. RDs are adjusted for age, sex, Charlson Comorbidity Index, educational level, and select pre-existing conditions (chronic diseases).

**Figure S4:**
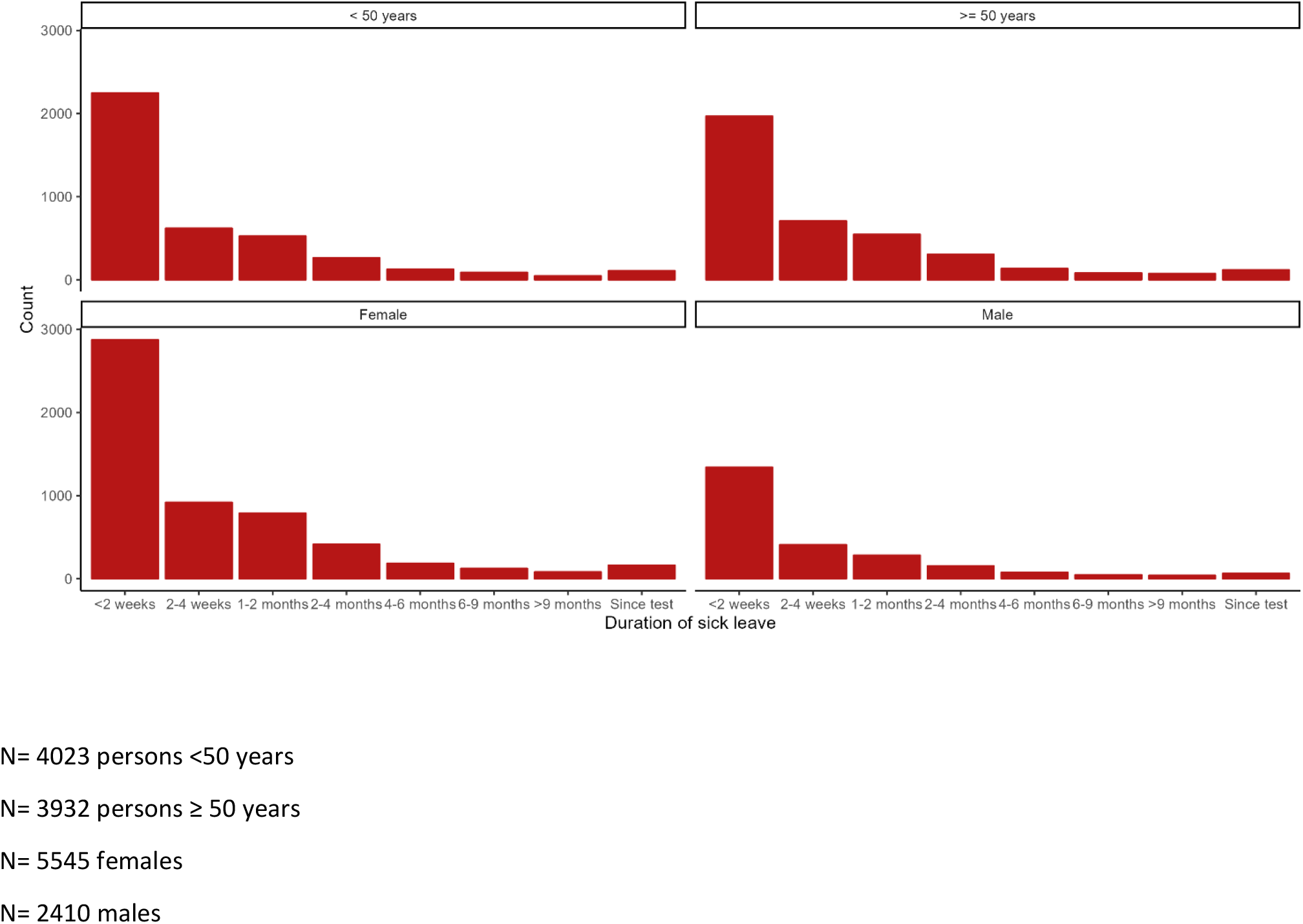
A distributional plot of the underlying self-reported sick leave by age (<50 years, ≥50 years) and sex.

**Table S1:**
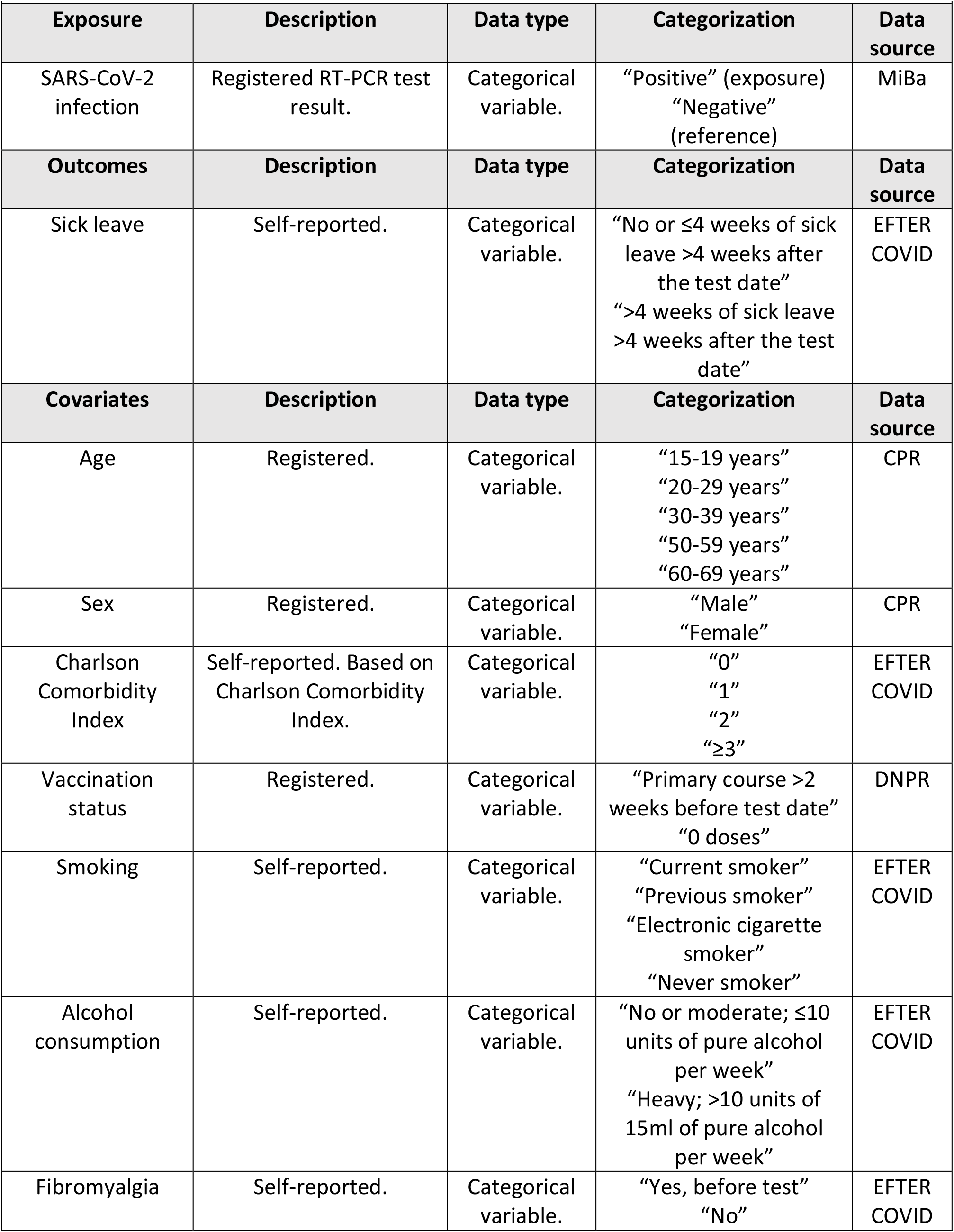
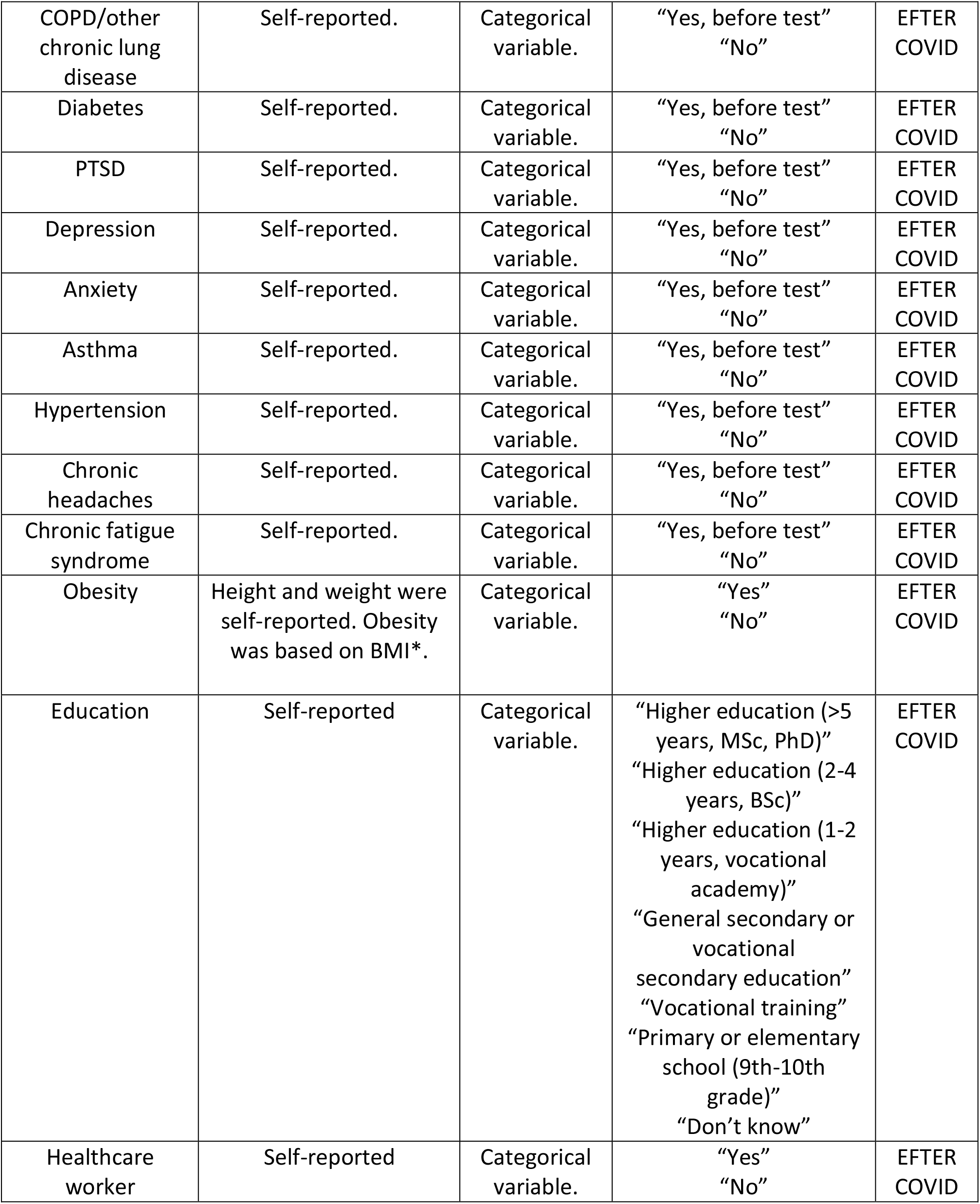
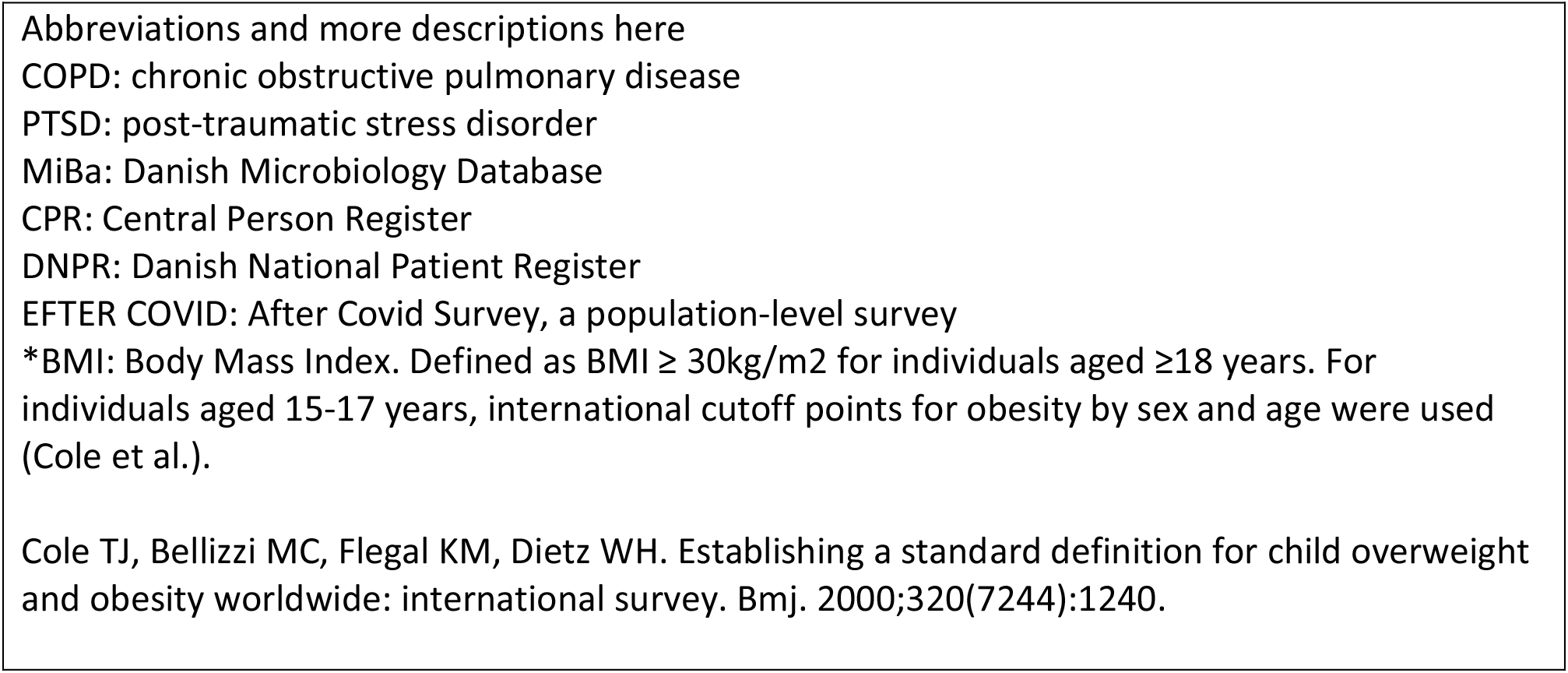
Overview of study variables and data sources

| **Exposure** | **Description** | **Data type** | **Categorization** | **Data source** |
| --- | --- | --- | --- | --- |
| SARS-CoV-2 infection | Registered RT-PCR test result. | Categorical variable. | “Positive” (exposure)<br>“Negative” (reference) | MiBa |
| **Outcomes** | **Description** | **Data type** | **Categorization** | **Data source** |
| Sick leave | Self-reported. | Categorical variable. | “No or ≤4 weeks of sick leave >4 weeks after the test date”<br>“>4 weeks of sick leave >4 weeks after the test date” | EFTER COVID |
| **Covariates** | **Description** | **Data type** | **Categorization** | **Data source** |
| Age | Registered. | Categorical variable. | “15-19 years”<br>“20-29 years”<br>“30-39 years”<br>“50-59 years”<br>“60-69 years” | CPR |
| Sex | Registered. | Categorical variable. | “Male”<br>“Female” | CPR |
| Charlson Comorbidity Index | Self-reported. Based on Charlson Comorbidity Index. | Categorical variable. | “0”<br>“1”<br>“2”<br>“≥3” | EFTER COVID |
| Vaccination status | Registered. | Categorical variable. | “Primary course >2 weeks before test date”<br>“0 doses” | DNPR |
| Smoking | Self-reported. | Categorical variable. | “Current smoker”<br>“Previous smoker”<br>“Electronic cigarette smoker”<br>“Never smoker” | EFTER COVID |
| Alcohol consumption | Self-reported. | Categorical variable. | “No or moderate; ≤10 units of pure alcohol per week”<br>“Heavy; >10 units of 15ml of pure alcohol per week” | EFTER COVID |
| Fibromyalgia | Self-reported. | Categorical variable. | “Yes, before test”<br>“No” | EFTER COVID |
| COPD/other chronic lung disease | Self-reported. | Categorical variable. | "Yes, before test"<br>"No" | EFTER COVID |
| Diabetes | Self-reported. | Categorical variable. | "Yes, before test"<br>"No" | EFTER COVID |
| PTSD | Self-reported. | Categorical variable. | "Yes, before test"<br>"No" | EFTER COVID |
| Depression | Self-reported. | Categorical variable. | "Yes, before test"<br>"No" | EFTER COVID |
| Anxiety | Self-reported. | Categorical variable. | "Yes, before test"<br>"No" | EFTER COVID |
| Asthma | Self-reported. | Categorical variable. | "Yes, before test"<br>"No" | EFTER COVID |
| Hypertension | Self-reported. | Categorical variable. | "Yes, before test"<br>"No" | EFTER COVID |
| Chronic headaches | Self-reported. | Categorical variable. | "Yes, before test"<br>"No" | EFTER COVID |
| Chronic fatigue syndrome | Self-reported. | Categorical variable. | "Yes, before test"<br>"No" | EFTER COVID |
| Obesity | Height and weight were self-reported. Obesity was based on BMI*. | Categorical variable. | "Yes"<br>"No" | EFTER COVID |
| Education | Self-reported | Categorical variable. | "Higher education (>5 years, MSc, PhD)"<br>"Higher education (2-4 years, BSc)"<br>"Higher education (1-2 years, vocational academy)"<br>"General secondary or vocational secondary education"<br>"Vocational training"<br>"Primary or elementary school (9th-10th grade)"<br>"Don't know" | EFTER COVID |
| Healthcare worker | Self-reported | Categorical variable. | "Yes"<br>"No" | EFTER COVID |

Abbreviations and more descriptions here
COPD: chronic obstructive pulmonary disease
PTSD: post-traumatic stress disorder
MiBa: Danish Microbiology Database
CPR: Central Person Register
DNPR: Danish National Patient Register
EFTER COVID: After Covid Survey, a population-level survey
\*BMI: Body Mass Index. Defined as $\text{BMI} \geq 30 \text{ kg/m}^2$ for individuals aged $\geq 18$ years. For individuals aged 15-17 years, international cutoff points for obesity by sex and age were used (Cole et al.).
Cole TJ, Bellizzi MC, Flegal KM, Dietz WH. Establishing a standard definition for child overweight and obesity worldwide: international survey. *Bmj*. 2000;320(7244):1240.

**Table S2:** Risk differences (RDs) and 95% confidence intervals (CI) for substantial sick leave taken one to nine months after the test date between SARS-CoV-2 test-negatives and positives for the total study population, stratified on educational level.

| Highest completed education | N | Sick leave among test-negatives | Sick leave among test-positives | RD (95% CI) |
| --- | --- | --- | --- | --- |
| Primary or elementary school (9<sup>th</sup>-10<sup>th</sup> grade) | 7258 | 48 (1.1%) | 122 (4.1%) | 3.7 (2.7, 4.7) |
| General secondary or vocational secondary education | 9838 | 58 (1.3%) | 116 (2.45) | 2.1 (1.3, 2.8) |
| Vocational training | 14245 | 142 (1.7%) | 318 (5.5%) | 3.9 (3.3, 4.6) |
| Higher education (1-2 years, vocational academy) | 10066 | 81 (1.3%) | 191 (4.7%) | 3.1 (2.5, 3.8) |
| Higher education (2-4 years, BSc) | 29384 | 272 (1.6%) | 673 (5.5%) | 3.8 (3.4, 4.2) |
| Higher education (>5 years, MSc, PhD) | 15737 | 115 (1.2%) | 207 (3.0%) | 1.9 (1.4, 2.4) |
| Don't know | 2289 | 16 (1.3 %) | 52 (5.1%) | 3.8 (2.2, 5.2) |
| N = 88,818 participants ages 15-65 years (response rate = 36%). N<sub>positive</sub> = 51,336, N<sub>negative</sub> = 37,482.<br>Risk differences are expressed in percentage points and adjusted for age, sex, Charlson Comorbidity Index, and select pre-existing conditions (chronic diseases). |  |  |  |  |

**Table S3:** Characteristics of test-positive population divided into subcategories based on substantial sick leave and long covid hospitalization.

| Background characteristics | Only positive test result<br>(N=35246) | Substantial sick leave<br>(N=1326) | Substantial sick leave and long covid hospitalization*<br>(N=354) | Only long covid hospitalization<br>(N=556) |
| --- | --- | --- | --- | --- |
| **Sex** |  |  |  |  |
| Female | 21390 (60.7%) | 956 (72.1%) | 261 (73.7%) | 395 (71.0%) |
| Male | 13856 (39.3%) | 370 (27.9%) | 93 (26.3%) | 161 (29.0%) |
| **Age** |  |  |  |  |
| Mean (SD) | 43.3 (14.0) | 48.4 (11.5) | 48.6 (11.2) | 46.7 (12.0) |
| Median [Min, Max] | 46.0 [15.0, 65.0] | 50.0 [15.0, 65.0] | 50.0 [20.0, 65.0] | 48.0 [15.0, 65.0] |
| **Charlson Comorbidity Index** |  |  |  |  |
| 0 | 32101 (91.1%) | 1118 (84.3%) | 289 (81.6%) | 480 (86.3%) |
| 1 | 1759 (5.0%) | 118 (8.9%) | 40 (11.3%) | 50 (9.0%) |
| 2 | 1038 (2.9%) | 66 (5.0%) | 18 (5.1%) | 22 (4.0%) |
| 3 or more | 348 (1.0%) | 24 (1.8%) | 7 (2.0%) | 4 (0.7%) |
| **Educational level (highest)** |  |  |  |  |
| Higher education (>5 years, MSc, PhD) | 6509 (18.5%) | 173 (13.0%) | 34 (9.6%) | 83 (14.9%) |
| Higher education (2-4 years, BSc) | 11235 (31.9%) | 502 (37.9%) | 171 (48.3%) | 239 (43.0%) |
| Higher education (1-2 years, vocational academy) | 3814 (10.8%) | 159 (12.0%) | 2 (9.0%) | 62 (11.2%) |
| Primary or elementary school (9<sup>th</sup>-10<sup>th</sup> grade) | 2838 (8.1%) | 102 (7.7%) | 20 (5.6%) | 31 (5.6%) |
| General secondary or vocational secondary education | (12.9%) | 98 (7.4%) | 18 (5.1%) | 37 (6.7%) |
| Vocational training | 5343 (15.2%) | 247 (18.6%) | 71 (20.1%) | 94 (16.9%) |
| Don't know | 949 (2.7%) | 44 (3.3%) | 8 (2.3%) | 10 (1.8%) |
| **Healthcare worker** |  |  |  |  |
| No | 30471 (86.5%) | 1097 (82.7%) | 266 (75.1%) | 450 (80.9%) |
| Yes | 4775 (13.5%) | 229 (17.3%) | 88 (24.9%) | 88 (24.9%) |
| **obesity** |  |  |  |  |
| no | 27146 (77.0%) | 850 (64.1%) | 213 (60.2%) | 390 (70.1%) |
| unknown | 2507 (7.1%) | 118 (8.9%) | 25 (7.1%) | 30 (5.4%) |
| yes | 5593 (15.9%) | 358 (27.0%) | 116 (32.8%) | 136 (24.5%) |
| **Fibromyalgia** |  |  |  |  |
| No | 34643 (98.3%) | 1270 (95.8%) | 330 (93.2%) | 540 (97.1%) |
| Before test | 285 (0.8%) | 30 (2.3%) | 9 (2.5%) | 5 (0.9%) |
| **Chronic fatigue syndrome** |  |  |  |  |
| No | 33671 (95.5%) | 1184 (89.3%) | 290 (81.9%) | 488 (87.8%) |
| Before test | 434 (1.2%) | 29 (2.2%) | 7 (2.0%) | 12 (2.2%) |
| **Anxiety** |  |  |  |  |
| No | 31495 (89.4%) | 1061 (80.0%) | 284 (80.2%) | 461 (82.9%) |
| Before test | 2765 (7.8%) | 155 (11.7%) | 21 (5.9%) | 53 (9.5%) |
| **Depression** |  |  |  |  |
| No | 30449 (86.4%) | 926 (69.8%) | 259 (73.2%) | 434 (78.1%) |
| Before test | 3840 (10.9%) | 277 (20.9%) | 53 (15.0%) | 79 (14.2%) |
| **Post traumatic stress disorder** |  |  |  |  |
| No | 34217 (97.1%) | 1236 (93.2%) | 324 (91.5%) | 520 (93.5%) |
| Before test | 636 (1.8%) | 49 (3.7%) | 12 (3.4%) | 23 (4.1%) |
| **Asthma** |  |  |  |  |
| No | 32693 (92.8%) | 1165 (87.9%) | 304 (85.9%) | 481 (86.5%) |
| Before test | 2553 (7.2%) | 161 (12.1%) | 50 (14.1%) | 75 (13.5%) |
| **Diabetes** |  |  |  |  |
| No | 34141 (96.9%) | 1248 (94.1%) | 330 (93.2%) | 536 (96.4%) |
| Before test | 1105 (3.1%) | 78 (5.9%) | 24 (6.8%) | 20 (3.6%) |
| **High blood pressure** |  |  |  |  |
| No | 31678 (89.9%) | 1119 (84.4%) | 299 (84.5%) | 493 (88.7%) |
| Before test | 3568 (10.1%) | 207 (15.6%) | 55 (15.5%) | 63 (11.3%) |
| **COPD or other lung disease** |  |  |  |  |
| No | 34868 (98.9%) | 1295 (97.7%) | 344 (97.2%) | 549 (98.7%) |
| Before test | 378 (1.1%) | 31 (2.3%) | 10 (2.8%) | 7 (1.3%) |
| **Chronic or frequent headaches, including migraines** |  |  |  |  |
| No | 33934 (96.3%) | 1227 (92.5%) | 330 (93.2%) | 522 (93.9%) |
| Before test | 1312 (3.7%) | 99 (7.5%) | 24 (6.8%) | 34 (6.1%) |
| *Long covid hospitalization is defined by the ICD-10 code B948A registered in the Danish National Patient Register (DNPR). |  |  |  |  |

## References

1. Crook H, Raza S, Nowell J, Young M, Edison P. Long covid—mechanisms, risk factors, and management. BMJ. 2021;374:n1648.

2. World Health Organization. Coronavirus disease (COVID-19): Post COVID-19 condition: World Health Organization; 2021 [Available from: https://www.who.int/news-room/questions-and-answers/item/coronavirus-disease-(covid-19)-post-covid-19-condition.

3. Taquet M, Geddes JR, Husain M, Luciano S, Harrison PJ. 6-month neurological and psychiatric outcomes in 236 379 survivors of COVID-19: a retrospective cohort study using electronic health records. Lancet Psychiatry. 2021;8(5):416–27.

4. Collaborators GBoDLC. Estimated Global Proportions of Individuals With Persistent Fatigue, Cognitive, and Respiratory Symptom Clusters Following Symptomatic COVID-19 in 2020 and 2021. JAMA. 2022;328(16):1604–15.

5. Peter RS, Nieters A, Kräusslich H-G, Brockmann SO, Göpel S, Kindle G, et al. Post-acute sequelae of covid-19 six to 12 months after infection: population based study. bmj. 2022;379.

6. Pfaff ER, Madlock-Brown C, Baratta JM, Bhatia A, Davis H, Girvin A, et al. Coding Long COVID: Characterizing a new disease through an ICD-10 lens. medRxiv. 2022:2022.04.18.22273968.

7. Davis HE, McCorkell L, Vogel JM, Topol EJ. Long COVID: major findings, mechanisms and recommendations. Nature Reviews Microbiology. 2023.

8. Mayor N, Meza-Torres B, Okusi C, Delanerolle G, Chapman M, Wang W, et al. Developing a Long COVID Phenotype for Postacute COVID-19 in a National Primary Care Sentinel Cohort: Observational Retrospective Database Analysis. JMIR Public Health and Surveillance. 2022;8(8):e36989.

9. Sudre CH, Murray B, Varsavsky T, Graham MS, Penfold RS, Bowyer RC, et al. Attributes and predictors of long COVID. Nature medicine. 2021;27(4):626–31.

10. Thompson EJ, Williams DM, Walker AJ, Mitchell RE, Niedzwiedz CL, Yang TC, et al. Risk factors for long COVID: analyses of 10 longitudinal studies and electronic health records in the UK. MedRxiv. 2021.

11. Tsampasian V, Elghazaly H, Chattopadhyay R, Debski M, Naing TKP, Garg P, et al. Risk Factors Associated With Post−COVID-19 Condition: A Systematic Review and Meta-analysis. JAMA Internal Medicine. 2023.

12. Davis HE, Assaf GS, McCorkell L, Wei H, Low RJ, Re’em Y, et al. Characterizing long COVID in an international cohort: 7 months of symptoms and their impact. EClinicalMedicine. 2021;38:101019.

13. Sørensen AIV, Spiliopoulos L, Bager P, Nielsen NM, Hansen JV, Koch A, et al. A nationwide questionnaire study of post-acute symptoms and health problems after SARS-CoV-2 infection in Denmark. Nature Communications. 2022;13(1):4213.

14. Jacob L, Koyanagi A, Smith L, Tanislav C, Konrad M, van der Beck S, et al. Prevalence of, and factors associated with, long-term COVID-19 sick leave in working-age patients followed in general practices in Germany. International Journal of Infectious Diseases. 2021;109:203–8.

15. Westerlind E, Palstam A, Sunnerhagen KS, Persson HC. Patterns and predictors of sick leave after Covid-19 and long Covid in a national Swedish cohort. BMC Public Health. 2021;21(1):1023.

16. Palstam A, Westerlind E, Sunnerhagen KS, Persson HC. Recurrent sick leave after COVID-19: investigating the first wave of the pandemic in a comprehensive Swedish registry-based study. BMC Public Health. 2021;21(1):1914.

17. Nielsen TB, Leth S, Pedersen M, Harbo HD, Nielsen CV, Laursen CH, et al. Mental Fatigue, Activities of Daily Living, Sick Leave and Functional Status among Patients with Long COVID: A Cross-Sectional Study. Int J Environ Res Public Health. 2022;19(22).

18. Hansen CH, Friis NU, Bager P, Stegger M, Fonager J, Fomsgaard A, et al. Risk of reinfection, vaccine protection, and severity of infection with the BA.5 omicron subvariant: a nation- wide population-based study in Denmark. Lancet Infect Dis. 2022.

19. European Centre for Disease Prevention and Control. Data on testing for COVID-19 by week and country: European Centre for Disease Prevention and Control,; 2022 [Available from: https://www.ecdc.europa.eu/en/publications-data/covid-19-testing.

20. Gram MA, Steenhard N, Cohen AS, Vangsted A-M, Mølbak K, Gorm Jensen T, et al. Patterns of testing in the extensive Danish national SARS-CoV-2 test set-up. medRxiv. 2023:2023.02.06.23285556.

21. Danish Ministry of Employment. Sick Leave: Agency for Digital Government, Ministry of Finance; 2022 [Available from: https://lifeindenmark.borger.dk/working/work-rights/leave-of-absence/sick-leave-and-sickness-benefit.

22. Digitaliseringsstyrelsen. Digitaliseringsstyrelsen; 2022 [Available from: https://digst.dk/.

23. SurveyXact. Ramboll; 2022 [Available from: https://www.surveyxact.dk/.

24. O’Regan E, Svalgaard I, Sørensen AIV, Spiliopoulos L, Bager P, Nielsen N, et al. Self-Reported Adverse Events Following SARS-CoV-2 Vaccination: A Nationwide Questionnaire Study in the Adult Danish Population. Available at SSRN 4231237.

25. Spiliopoulos L, Sørensen AIV, Bager PM, Nielsen NM, Hansen JV, Koch A, et al. Post-acute symptoms four months after SARS-CoV-2 infection during the Omicron period: a nationwide Danish questionnaire study. medRxiv. 2022.

26. Statens Serum Institut. Næsten en million danskere har bidraget med viden om senfølger efter covid-19: Statens Serum Institut; 2023 [Available from: https://www.ssi.dk/aktuelt/nyheder/2023/naesten-en-million-danskere-har-bidraget-med-viden-om-senfolger-efter-covid19.

27. Robins J. A new approach to causal inference in mortality studies with a sustained exposure period—application to control of the healthy worker survivor effect. Mathematical Modelling. 1986;7(9):1393–512.

28. Team RC. R: A language and environment for statistical computing. R Foundation for Statistical Computing, Vienna, Austria. http://www.R-project.org/. 2013.

29. Grembi J ME. riskCommunicator: G-Computation to Estimate Interpretable Epidemiological Effects. R package version 1.0.1 2020. [Available from: https://cran.r-project.org/web/packages/riskCommunicator/index.html.

30. Alimu D. forestploter; Create Flexibile Forest Plot. R package version 0.1.6. . 2022.

31. Westerlind E, Palstam A, Sunnerhagen KS, Persson HC. Patterns and predictors of sick leave after Covid-19 and long Covid in a national Swedish cohort. BMC public health. 2021;21(1):1–9.

32. Jacobsen PA, Andersen MP, Gislason G, Phelps M, Butt JH, Køber L, et al. Return to work after COVID-19 infection - A Danish nationwide registry study. Public Health. 2022;203:116–22.

33. Skyrud K, Telle K, Magnusson K. Impacts of mild and severe COVID-19 on sick leave. Int J Epidemiol. 2021;50(5):1745–7.

34. European Centre for Disease Prevention and Control. Covid-19 / Latest evidence / Risk factors and risk groups: European Centre for Disease Prevention and Control,; 2022 [Available from: https://www.ecdc.europa.eu/en/covid-19/latest-evidence/risk-factors-risk-groups.

35. Russell CD, Lone NI, Baillie JK. Comorbidities, multimorbidity and COVID-19. Nature Medicine. 2023;29(2):334–43.

36. Ghosh P, Niesen MJM, Pawlowski C, Bandi H, Yoo U, Lenehan PJ, et al. Severe acute infection and chronic pulmonary disease are risk factors for developing post-COVID-19 conditions. medRxiv. 2022.

37. Grøsland M, Reme BA, Gjefsen HM. Impact of Omicron on sick leave across industries: A population-wide study. Scand J Public Health. 2022:14034948221123163.

38. Dencker-Larsen S, Rasmussen CL, Thorsen SV, Clays E, Lund T, Labriola M, et al. Technically measured compositional physical work demands and prospective register-based sickness absence (PODESA): a study protocol. BMC Public Health. 2019;19(1):257.

